# Somatic mutations in chronic lung disease are associated with reduced lung function

**DOI:** 10.1101/2023.03.03.23286771

**Authors:** Jeong H. Yun, M.A. Wasay Khan, Auyon Ghosh, Brian D. Hobbs, Peter J. Castaldi, Craig P. Hersh, Peter G. Miller, Carlyne D. Cool, Frank Sciurba, Lucas Barwick, Andrew H. Limper, Kevin Flaherty, Gerard J. Criner, Kevin Brown, Robert Wise, Fernando Martinez, Edwin K. Silverman, Dawn Demeo, Michael H. Cho, Alexander G. Bick

**Affiliations:** Channing Division of Network Medicine, Department of Medicine, Brigham and Women’s Hospital; Division of Pulmonary and Critical Care Medicine, Department of Medicine, Brigham and Women’s Hospital; Harvard Medical School; Division of Genetic Medicine, Department of Medicine, Vanderbilt University; Pulmonary Critical Care and Sleep Medicine, Upstate Medical University; Center for Cancer Research, Massachusetts General Hospital; Division of Pathology, Department of Medicine, National Jewish Health; Division of Pulmonary, Allergy and Critical Care Medicine, University of Pittsburgh; Emmes; Division of Pulmonary and Critical Care Medicine, Department of Internal Medicine, Mayo Clinic; Division of Pulmonary & Critical Care Medicine, University of Michigan Health System; Thoracic Medicine and Surgery, Lewis Katz School of Medicine at Temple University; Department of Medicine, National Jewish Health; Department of Medicine, Johns Hopkins Medicine; Department of Medicine, Weill Cornell Medical College

## Abstract

Among human organs, the lung harbors one of the highest rates of somatic mutations. However, the relationship of these mutations to lung disease and function is not known. We analyzed the somatic mutational pattern from 1,251 samples of normal and diseased non-cancerous lung tissue from the Lung Tissue Research Consortium using RNA-seq. In two of the most common diseases represented in our dataset, chronic obstructive pulmonary disease (COPD, 29%) and idiopathic pulmonary fibrosis (IPF, 13%), we found a significantly increased burden of somatic mutations compared to normal. Using deconvoluted cell type proportions, we found that a major predictor of somatic mutations was the airway to alveolar cell proportion and pathogenic cell types. We also found that mutational burden was associated with reduced lung function. This relationship remained even after adjustment for age, sex, smoking, and cell type proportion and in COPD and IPF. Our identification of an increased prevalence of somatic mutation in diseased lung that correlates with cell type proportion and disease severity highlights for the first time the role of somatic mutational processes in lung disease genetics.

## Main

Lung tissue has one of the highest somatic mutation rates across normal tissues^1^. The contribution of the somatic mutations from tobacco smoking leading to lung cancer is well established, and studies have shown that tobacco smoking increases the mutational burden of normal human bronchial epithelial cells^2,3^. However, while acquired somatic mutations have been proposed as contributors to disease pathogenesis^4–6^, the relationship of mutational burden to lung function and disease has not been well studied.

We examined the patterns of somatic single nucleotide variants (sSNVs) in lung tissue using bulk RNA-seq from lung specimens in 1,251 subjects in the Lung Tissue Research Consortium (LTRC) from the Trans-Omics for Precision Medicine program^7^. LTRC lung tissue samples were collected on well characterized individuals who underwent clinically indicated thoracic surgery, including lung transplantation, lung volume reduction surgery for emphysema, or pulmonary nodule and cancer resection. The majority of subjects (65%) had chronic lung diseases; the most common of these were COPD and idiopathic pulmonary fibrosis (IPF) (Supplementary Table 1). Four hundred seventeen (33%) subjects had lung cancer at the time of surgery. Lung tissue samples were isolated from regions without evidence of possible lung cancer and reviewed by the LTRC pathology core.

We applied the RNA-MuTect pipeline^1^ to lung tissue RNA-seq data and performed cellular deconvolution of cell type categories^8^. RNA-MuTect has been validated across multiple tissue types, including the lung, to detect DNA mutations with high allele fractions (>7%) in the RNA_1_. To avoid misclassification of germline genetic mutations as somatic mutations from the RNA-seq data, we excluded germline mutations identified using whole-genome DNA sequencing from paired blood samples and common variants with median allele frequency (MAF) greater than 30%. To avoid recurrent sequencing artifacts, we removed mutations that were observed in more than 10 individuals. To ensure that we had high confidence in the somatic mutations, we removed variants with MAF <10% and variants with fewer than 2 supporting reads. We also removed individuals who had a total number of mutations that were outside the interquartile range, resulting in 113,657 high confidence coding region mutations in 1,251 individuals (Fig 1). A separate analysis was performed in subgroups with smoking associated chronic lung diseases, COPD (n=358), IPF (n=163) and normal controls (n=29), the latter three groups confirmed by histopathology and lung function tests (Fig 1, Supplementary Table 2).

**Fig 1.**
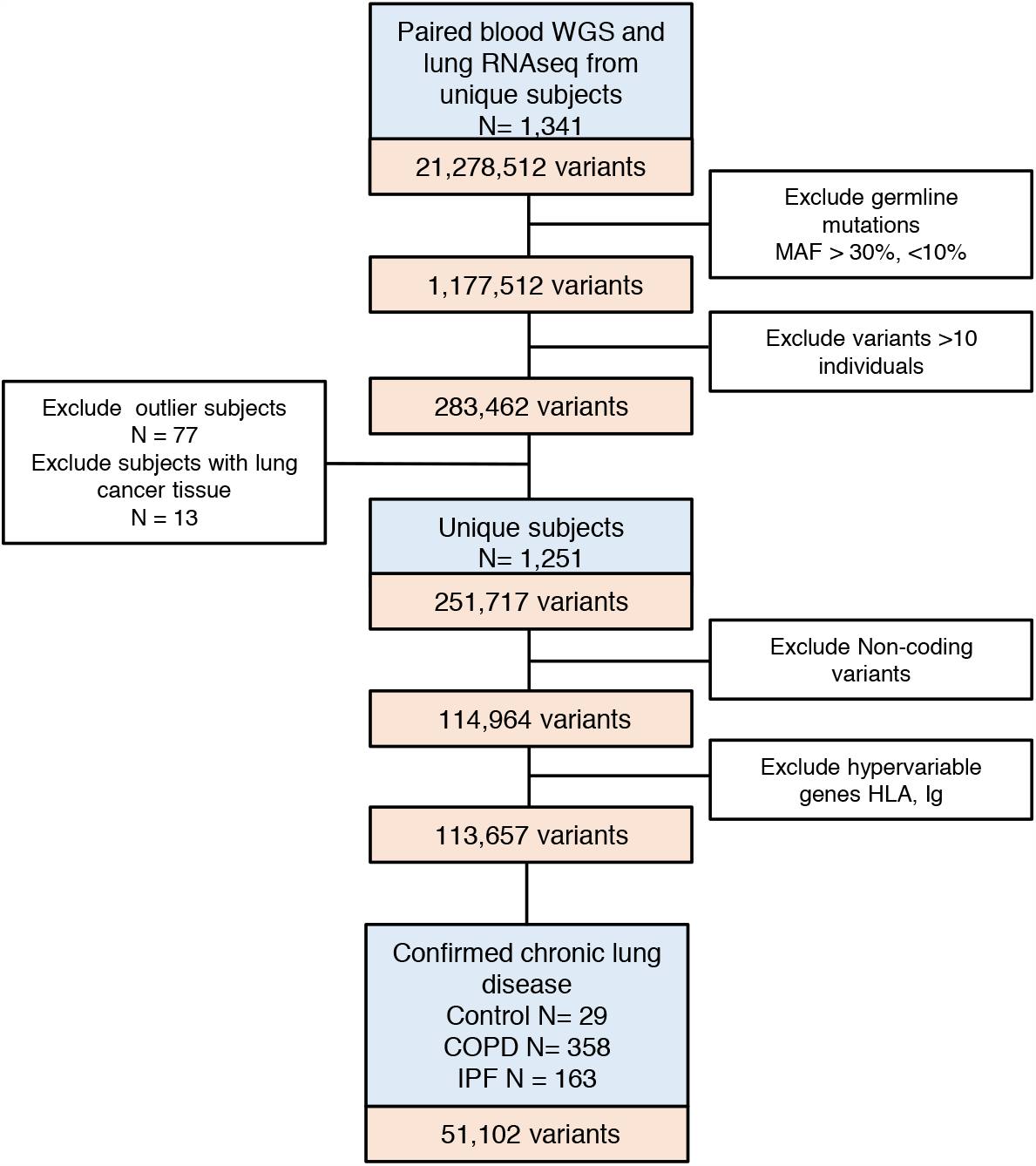
Study design. Abbreviations: WGS whole genome sequencing, MAF major allele frequency, COPD chronic obstructive pulmonary disease, IPF idiopathic pulmonary fibrosis

Among 1,251 subjects, missense mutations (52%) were more common than silent mutations (47%). The most common SNV classes were C>T (47%) and T>C (26%) mutations. The median number of sSNVs per sample was 87 (46 excluding silent mutation) (Extended Data Fig 1).

Somatic mutation frequency linearly increases with age in normal human bronchial epithelial cells, and at a higher rate in smokers^3^. In our data, we did not find a significant overall association between mutational burden (total number of sSNVs) and chronological age, although a trend towards positive association with age was observed in the control group (Extended data Fig 2, Fig 2). Similarly, we found a weak inverse association with cumulative smoking (τ = -0.066, p= 0.001). However, we found that the overall mutational burden was significantly elevated in COPD and IPF patients compared to controls (Supplementary Table 2) as well as to published data of normal lung tissue using the same method^1^. LTRC subjects overall have a high cumulative tobacco smoke exposure in the range where the mutation frequencies level off^3^. Thus, we postulate that due to high baseline mutational burden in the diseased tissue and the significant smoking history across the cohort, age and cumulative smoking history do not correlate with mutational burden as has been shown in populations without such constraints^3^.

**Fig 2.**
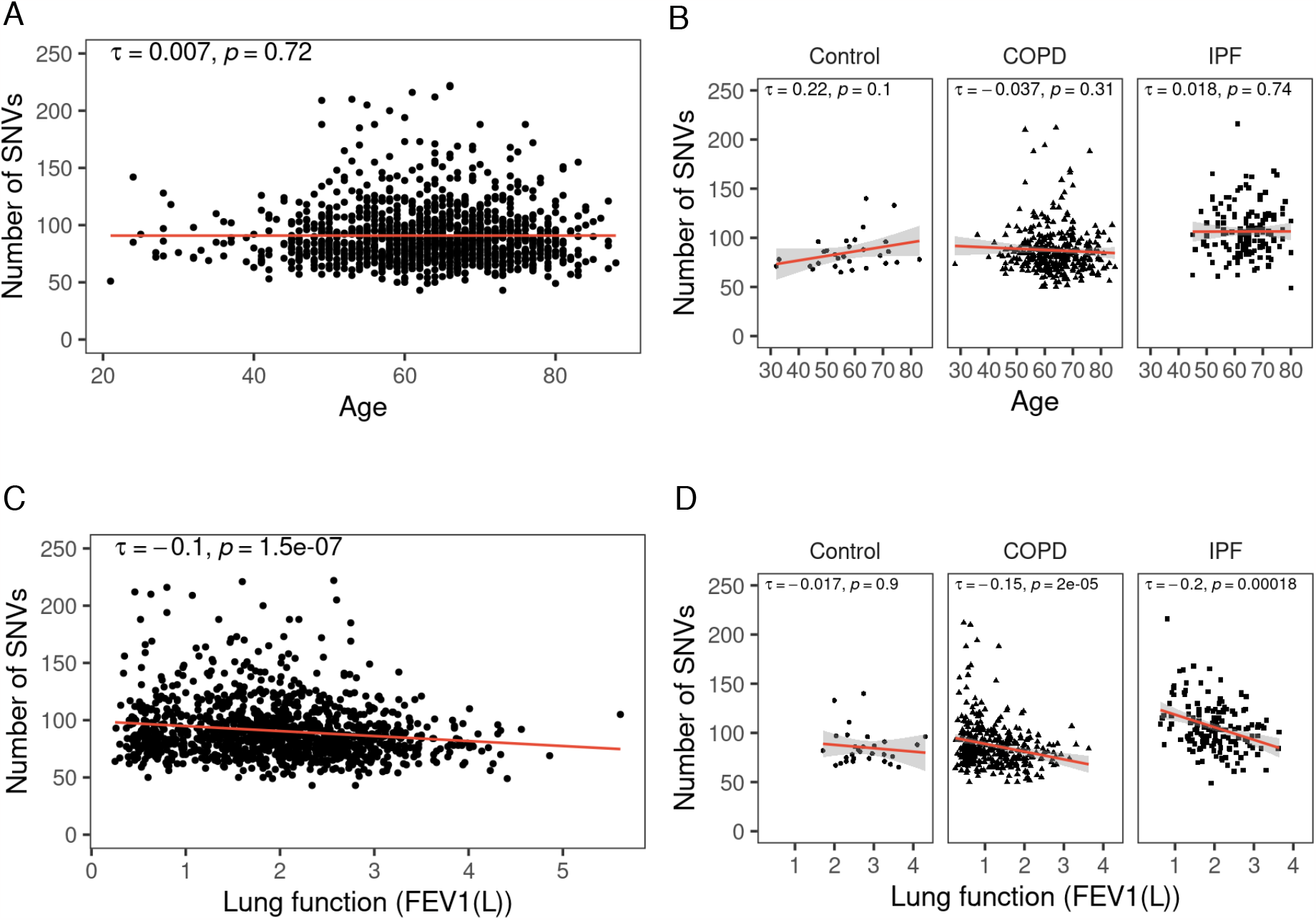
Mutational burden is inversely associated with lung function in COPD and IPF. A. Lack of association between number of SNVs and age in total subjects or B. subgroups. C. Lung function (FEV_1_(L)) is inversely associated with age in total subects and in B. COPD and IPF. Kendall rank correlation coefficient shown. Abbreviations: SNV single nucleotide variant, COPD chronic obstructive pulmonary disease, IPF idiopathic pulmonary fibrosis, FEV_1_ forced expiratory volume (L) in 1 second.

We next investigated mutational burden was associated with metrics of disease severity. Lung function, such as forced expiratory volume in one second (FEV_1_) and forced vital capacity (FVC), is a physiologic measure that assess the integrated function of the respiratory system. Lung function decline normally begins after age 35 and accelerates with smoking or lung disease processes^9^, reflecting disease severity. We detected a strong inverse association between mutational burden and lung function, as assessed by both FEV_1_ (τ -0.1, p = 1.5x 10^−7^) and FVC (τ -0.16, p <2.2x 10^−16^) (Fig 2, Extended Data Fig 3). In a multivariable linear regression model to control for known determinants of lung function including age, sex, height, race and smoking history, and found that the lung somatic mutational burden remained a statistically significant predictor of lung function in all subjects, as well as in the disease subgroups (Table 1, Supplementary Table 3).

**Table 1.**
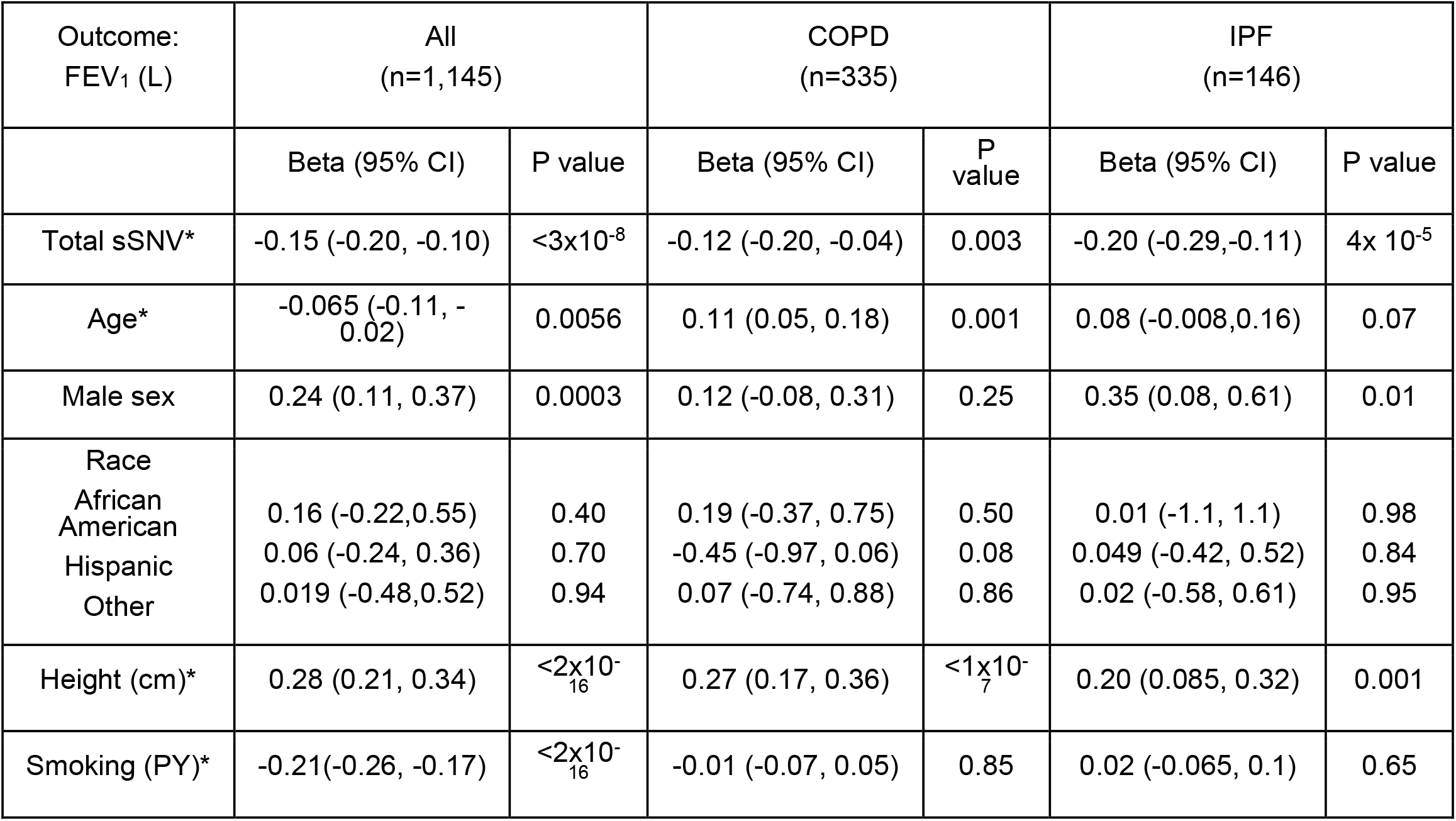
Lung function (FEV_1_ (L)) is associated with lung somatic mutational burden. Multivariable linear regression. *Continuous variables are standardized. FEV_1_ forced expiratory volume in 1 second. COPD chronic obstructive pulmonary disease, IPF idiopathic pulmonary fibrosis. sSNV somatic single nucleotide variant, PY pack-years

| Outcome:<br>FEV<sub>1</sub> (L) | All<br>(n=1,145) |  | COPD<br>(n=335) |  | IPF<br>(n=146) |  |
| --- | --- | --- | --- | --- | --- | --- |
|  | Beta (95% CI) | P value | Beta (95% CI) | P value | Beta (95% CI) | P value |
| Total sSNV* | -0.15 (-0.20, -0.10) | <3x10<sup>-8</sup> | -0.12 (-0.20, -0.04) | 0.003 | -0.20 (-0.29,-0.11) | 4x 10<sup>-5</sup> |
| Age* | -0.065 (-0.11, -0.02) | 0.0056 | 0.11 (0.05, 0.18) | 0.001 | 0.08 (-0.008,0.16) | 0.07 |
| Male sex | 0.24 (0.11, 0.37) | 0.0003 | 0.12 (-0.08, 0.31) | 0.25 | 0.35 (0.08, 0.61) | 0.01 |
| Race |  |  |  |  |  |  |
| African American | 0.16 (-0.22,0.55) | 0.40 | 0.19 (-0.37, 0.75) | 0.50 | 0.01 (-1.1, 1.1) | 0.98 |
| Hispanic | 0.06 (-0.24, 0.36) | 0.70 | -0.45 (-0.97, 0.06) | 0.08 | 0.049 (-0.42, 0.52) | 0.84 |
| Other | 0.019 (-0.48,0.52) | 0.94 | 0.07 (-0.74, 0.88) | 0.86 | 0.02 (-0.58, 0.61) | 0.95 |
| Height (cm)* | 0.28 (0.21, 0.34) | <2x10<sup>-16</sup> | 0.27 (0.17, 0.36) | <1x10<sup>-7</sup> | 0.20 (0.085, 0.32) | 0.001 |
| Smoking (PY)* | -0.21(-0.26, -0.17) | <2x10<sup>-16</sup> | -0.01 (-0.07, 0.05) | 0.85 | 0.02 (-0.065, 0.1) | 0.65 |

**Fig 3.**
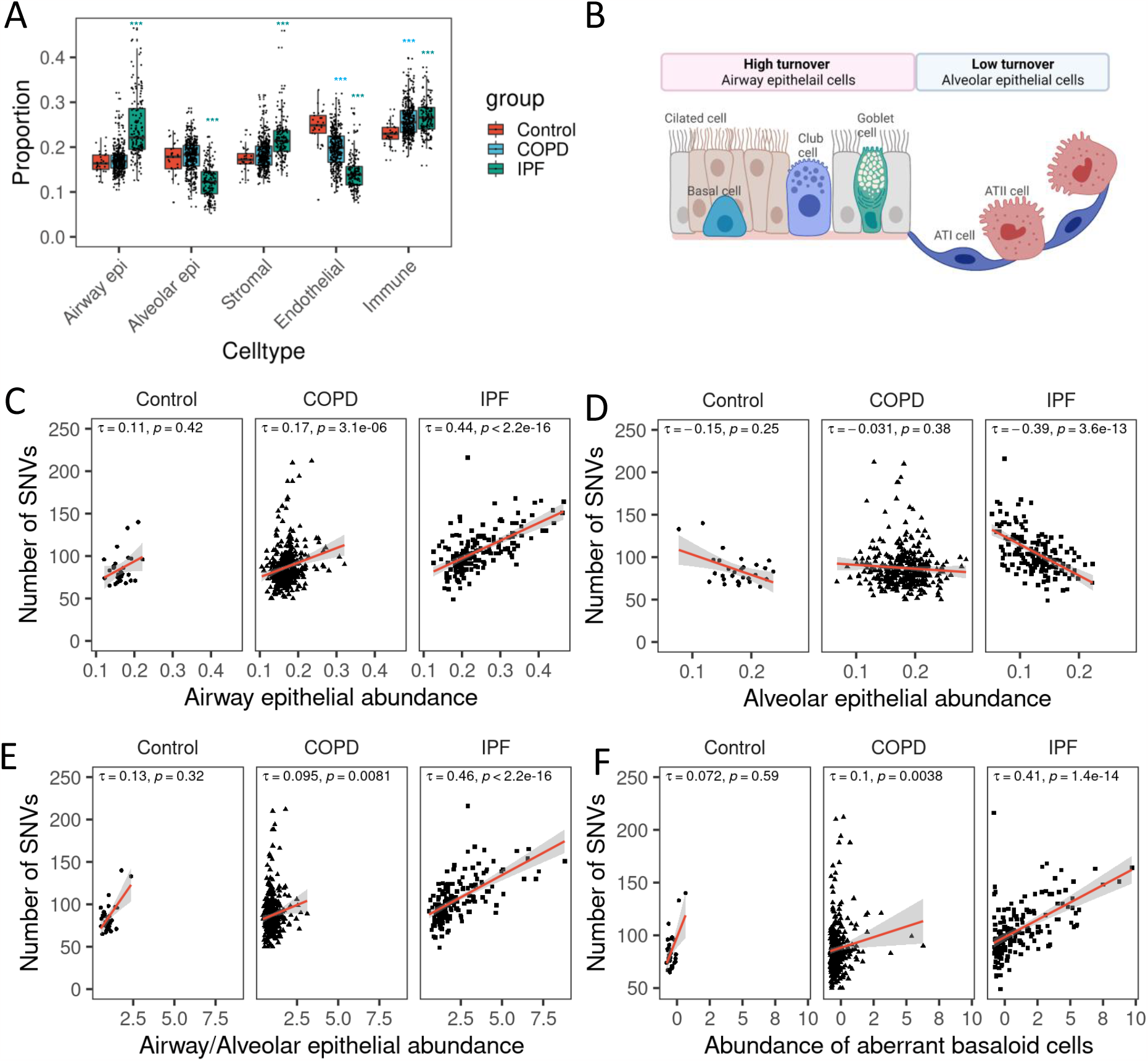
Mutational burden is associated with the proportion of high turnover cell types. A. Relative abundance of deconvoluted cell types from Bisque. *** t test adjusted p value <0.001 compared to controls. B. Schematic of airway and alveolar epithelial cell types. C-F. Correlation of somatic mutational burden and deconvoluted cell types. Aberrant basaloid cell was separately deconvoluted and shows relative abundance, not absolute proportions. Kendall rank correlation coefficient shown. Abbreviations: AT1 alveolar type 1 epithelial cells, AT2 alveolar type 2 epithelial cells, SNV single nucleotide variant, COPD chronic obstructive pulmonary disease, IPF idiopathic pulmonary fibrosis.

Lung tissue consists of heterogeneous cell types, and distinct disease processes can affect cell types differently. COPD is characterized by infiltration of inflammatory cells and loss of alveolar epithelial cells (emphysema)^10^, while IPF has progressive interstitial fibrosis with profibrotic macrophages and aberrant basal like cells^11,12^. We estimated the relative abundance of these cell types in each sample with the RNA expression data and examined how the cellular composition in the diseased lung tissue was associated with mutational burden. The relative abundance of airway epithelial cells, alveolar epithelial cells, stromal, immune and endothelial cells were quantified by deconvolution of RNA-seq data using Bisque^8^ as previously described^13^, and pathologic aberrant basaloid cells were separately deconvoluted with Bisque^8^ using a published IPF single cell RNA-seq reference dataset (Supplementary Table 4)^11^. Mutational burden was positively correlated with the proportion of airway epithelial cells and inversely correlated with the proportion of alveolar epithelial cells (Fig 3). The mutational burden was not significantly associated with stromal cells but was significantly associated with immune and endothelial cells in the IPF samples, possibly due to cellular proportion changes associated with IPF (Extended Data Fig 4). This pattern suggests that high turnover cells, such as airway epithelial cells, have higher accumulation of somatic mutations compared to low turnover cells, such as alveolar epithelial cells^14^. It is also notable that aberrant basaloid cells, most commonly found in IPF^11^, are captured within airway epithelial cells and their abundance correlated with mutational burden in both COPD and IPF (Fig 3). In multivariable regression analysis adjusted for age, sex, race, smoking history and total mapped reads, both airway/alveolar epithelial ratio and lung function (FEV_1,_ FVC) were statistically significantly and independently associated with mutational burden (Table 2, Supplementary Table 5).

**Table 2.**
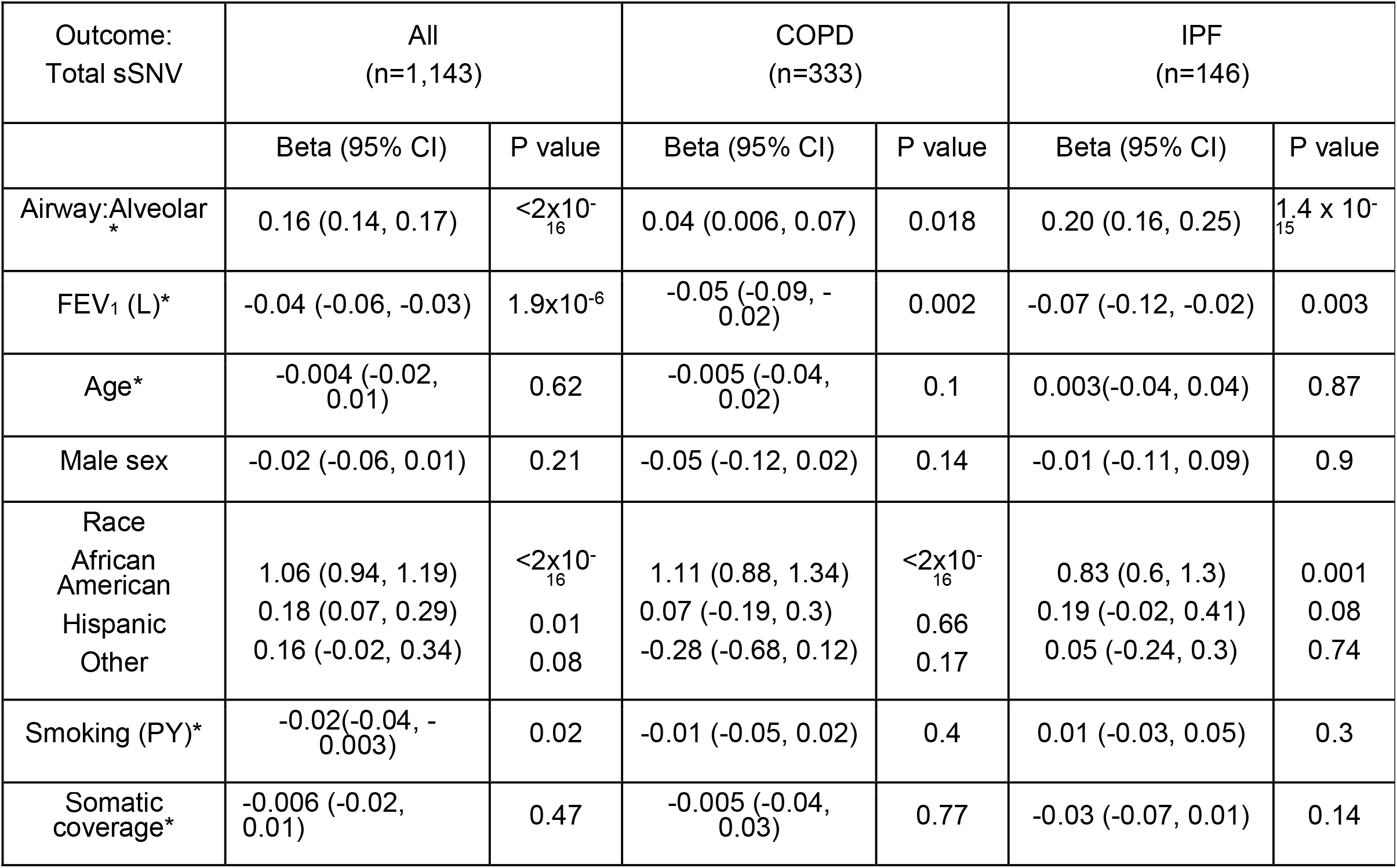
Lung somatic mutation burden is associated with lung function (FEV_1_) and airway to alveolar epithelial ratio. The number of total sSNV was log-transformed.

We observe that the most commonly mutated genes encoded for mucin proteins, which have elevated expression levels in IPF and advanced COPD patients^15^, and are also frequently mutated in public exomes (FLAGS)^16^. To rule out the possibility that differential gene expression, and thus sensitivity of detection by RNA-seq, was the primary driver the association between mutational burden, lung function, and cell type proportions, we performed two sensitivity analyses. First, we excluded the top 20 FLAGS genes (Supplementary Table 6) and confirmed that the relationship among mutational burden, cell types and lung function held (Extended Data Fig 5). Second, the number of sSNVs after normalizing by the length scaled gene expression level, the mutational burden remains elevated in IPF subjects (Extended Data Fig 6).

Utilizing large scale sequencing data with detailed clinical and pathologic phenotyping, we found that lung samples from patients with COPD and IPF lungs harbor a higher mutational burden compared to lung samples from normal controls. Although the detection of somatic variants from RNA-seq data is limited to macroscopic clones, by comparing different disease groups, we identified distinct and pervasive mutational patterns among chronic lung diseases and normal lungs. Our observations permit several conclusions.

First, we found the proliferative cell type, represented by the ratio of airway epithelial cells to alveolar epithelial cells, was strongly correlated with mutational burden across all samples, as well as in COPD and IPF. As mutations accumulate with each normal stem cell division, the tissues with a higher proliferation rate have higher number of mutations and increased cancer risk^19^. By inferring cell types within lung tissue, we extend this finding to cell types with different proliferation rates, as the number of cell divisions are one of the major cause of base substitution mutations from replication errors.

The proportion of airway to alveolar epithelial cells and its different strength of association with somatic mutational burden by disease states may also be explained by the cellular response in the diseased tissue. During homeostasis the airway and alveolar compartments are maintained by local progenitor populations such as basal cells in the airway and alveolar type 2 cells in alveolus. Lung regenerative process after injury or diseases are more complex and context specific, as different cell types are involved in distinct injury and repair process^20^. COPD is characterized by loss of alveolar epithelium from defective alveolar epithelial repair, while IPF is associated with aberrant repair by alveolar type 2 cells leading to accumulation of aberrant cells resembling basal cells^21^ that is captured as airway epithelium in current analysis. Therefore, the degree of defective or aberrant repair will reflect increased cellular turnover and be associated with increased airway to alveolar epithelial proportions.

Second, we also found a relationship between increased somatic mutation and reduced lung function. The independent association of lung function and mutational burden, despite the differences in cell types, composition and transcript levels implies that the function of the organ, the proximal correlate of disease severity, is closely related to the somatic mutational burden. The role of somatic mutations in the pathogenesis of COPD and IPF have long been hypothesized as both diseases are strongly associated tobacco smoking with evidence of DNA damage such as microsatellite instability and loss of heterozygosity have been observed^4–6,22^. Whether these somatic variants directly drive disease pathology remains to be determined given our cross-sectional design. However, even if these cells with clonal mutations arise via a selective advantage in the context of injured and inflamed tissue, their correlation with pathologic cell types and disease severity, suggest somatic mutations may contribute to disease progression, prognosis and risk of future malignancy, which is elevated in COPD and IPF independently from age and tobacco smoking and correlates with more severe disease^23^.

Our findings are similar to somatic mutational patterns reported in liver cirrhosis, where the number of somatic mutations correlated with known biomarkers of liver function but not with age or smoking status^24^. Future investigations on the association of somatic mutation frequency with the function of the organs in other disease contexts, such as glomerular filtration rate in the kidney or ejection fraction of the heart would be helpful to understand the global implication of somatic mutations in chronic non-malignant diseases. Our study is limited by cross-sectional design, lack of validation samples and reduced sensitivity relying on RNA sequencing data. Functional studies interrogating the impact of mutated genes would be required to determine the exact role of somatic mutations in COPD and IPF. Finally, the evaluation of somatic mutations from anatomically resolved tissue, or multiple different single cell types would further advance our understanding of how somatic mutations evolve and contribute to the pathology of chronic respiratory diseases, resulting from the complex interplay of genetic susceptibility, environmental exposures and aging.

## Data Availability

Data are available on the NCBI database of Genotypes and Phenotypes (dbGaP), accession phs001662

