## Supplementary Text for "Somatic mutations in chronic lung disease are associated with reduced lung function"

### Supplementary Materials

Supplementary Results

Supplementary Table 1.

Supplementary Table 2.

Supplementary Table 3.

Supplementary Table 4.

Supplementary Table 5.

Supplementary Table 6.

### Supplementary Results

In addition to sharing the risk factor of tobacco smoking, COPD and IPF patients have an increased risk of developing lung cancer, independent of smoking exposure[17,18]. To determine whether somatic mutations in chronic lung diseases are enriched for known lung cancer and cancer-driver genes (Supplementary File 1)[2], we compared the lung somatic mutational patterns by history of lung cancer, smoking history and disease states. Overall, somatic mutations in lung cancer driver genes were identified in 707 (56%) subjects. There was no significant enrichment of somatic mutations in known lung cancer driver genes in subgroups of individuals with cancer identified at the time of lung tissue collection or in stratum of smoking history (Extended Data Fig 7).

Next, we examined whether there are mutational signatures associated with aging, smoking or lung cancer in our data. We identified 2 single-base substitution signatures (SBS) from the entire set of subjects that are similar to SBS 6 (defective DNA mismatch repair) and SBS 25 (chemotherapy treatment). The normal controls and COPD subjects had similar signature profiles while IPF subjects had an additional signature related to clock-like signatures SBS 1 and 5 (Extended Data Fig 8).

| | No cancer (n=834) | Cancer (n=417) | P value |
|---|---|---|---|
| Age (mean(SD)) | 61.4 (10.8) | 67.1 (9.3) | <0.001 |
| Male (%) | 444 (53.2) | 216 (51.8) | 0.67 |
| Smoking history (%) | | | <0.001 |
| Never | 199 (23.9) | 52 (12.5) | |
| Former | 539 (64.6) | 300 (71.9) | |
| Current | 36 (4.3) | 32 (7.7) | |
| Packyears of smoking (mean (SD)) | 27.1 (29.4) | 40.6 (36.5) | <0.001 |
| FEV1 % predicted (mean(SD)) | 64.3 (28.2) | 75.2 (23) | <0.001 |
| Pathologic diagnosis | | | <0.001 |
| Normal | 29 (3.5) | 28 (6.7) | |
| Bronchiolitis | 55 (6.6) | 36 (8.6) | |
| Emphysema | 375 (45) | 282 (67.6) | |
| Fibrosis | 220 (26.4) | 23 (5.5) | |
| Granuloma | 27 (3.2) | 18 (4.3) | |
| Non-diagnostic | 15 (1.8) | 2 (0.5) | |
| Other* | 113 (13.5) | 28 (6.7) | |
| Clinical diagnosis (%) | | | <0.001 |
| Normal | 17 (2) | 0 | |
| Autoimmune | 19 (2.3) | 0 | |
| Bronchiolitis | 44 (5.3) | 0 | |
| Emphysema | 326 (39.1) | 0 | |
| Granulomatous infection | 54 (6.5) | 0 | |
| HP | 32 (3.8) | 0 | |
| NSCLC | 0 | 409 (98.1) | |
| SCLC | 0 | 8 (1.9) | |
| UIP | 196 (23.5) | 0 | |
| Other ILD** | 53 (6.4) | 0 | |
| Sarcoid | 13 (1.6) | 0 | |
| Lymphoma | 6 (0.7) | 0 | |
| Other | 74 (8.9) | 0 | |
| Somatic coverage (mean(SD)) | $1.89 \times 10^7$ ($2 \times 10^6$) | $1.87 \times 10^7$($2.2 \times 10^6$) | 0.17 |
| Total sSNV (mean(SD)) | 92.5 (24.7) | 87.6 (23.7) | 0.001 |
| Any cancer driver gene mutation (%) | 485 (58.2) | 241 (57.8) | 0.95 |

**Supplementary Table 1. Study Subjects**

*Central pathology diagnosis from LTRC tissue core laboratory.

**Includes cryptogenic organizing pneumonia, nonspecific interstitial pneumonia and uncharacterized fibrosis.

Abbreviations: $FEV_1$ forced expiratory volume in 1 second, BMI body mass index, HP hypersensitivity pneumonitis, NSCLC non-small cell lung cancer, SCLC small cell lung cancer, UIP usual interstitial pneumonia, ILD interstitial lung disease, sSNV somatic single nucleotide variant

| | Control (n=29) | COPD (n=358) | IPF (n = 163) | P value |
|---|---|---|---|---|
| Age (mean(SD)) | 57.9 (12.1) | 64.1 (9.0) | 64.1 (7.5) | 0.001 |
| Male (%) | 7 (24.1) | 185 (51.7) | 117 (71.8) | <0.001 |
| Current smokers (%) | 0 | 21 (6.2) | 2 (1.3) | 0.03 |
| Packyears of smoking (mean (SD)) | 8.0 (11.8) | 47.9 (30.8) | 19.3 (22.2) | <0.001 |
| FEV1 % predicted (mean(SD)) | 97.1 (10.6) | 42.9 (21.2) | 65.7 (19.5) | <0.001 |
| Emphysema (LAA950) mean(SD)) | 5.6 (5.4) | 23.1 (15.5) | 5.3 (6.3) | <0.001 |
| Pathologic diagnosis | | | | |
| Normal | 29 (100%) | 0 | 0 | |
| Emphysema | 0 | 358 (100%) | 0 | |
| UIP/honeycombing | 0 | 0 | 163 (100%) | |
| Clinical diagnosis (%) | | | | |
| Normal | 7 (24.1) | 0 | 0 | |
| Autoimmune | 0 | 0 | 4 (2.5) | |
| Bronchiolitis | 0 | 3 (0.8) | 1 (0.6) | |
| Emphysema | 0 | 198 (55.3) | 4 (2.5) | |
| Uncharacterized fibrosis | 0 | 2 (0.6) | 2 (1.2) | |
| Granuloma infection | 0 | 15 (4.2) | 0 | |
| HP | 0 | 0 | 8 (4.9) | |
| NSCLC | 15 (51.7) | 130 (36.3) | 6 (3.7) | |
| SCLC | 6 (20.7) | 2 (0.6) | 0 | |
| UIP | 0 | 0 | 135 (82.8) | |
| Other | 0 | 8 (2.2) | 3 (1.8) | |
| Somatic coverage (mean(SD)) | $1.89 \times 10^7$ ($1.9 \times 10^6$) | $1.87 \times 10^7$ ($2.3 \times 10^6$) | $1.89 \times 10^7$($2.3 \times 10^6$) | 0.67 |
| Total aSNV (mean(SD)) | 85.2 (18) | 87.2 (22.9) | 106.4 (24.4) | <0.001 |
| Any cancer driver gene mutation (%) | 11 (37.9) | 219 (61.2) | 93 (57.1) | 0.04 |

**Supplementary Table 2. Normal, COPD and IPF subjects**

Control subjects had normal pathology and normal lung function, COPD subjects were defined by moderate to severe airflow obstruction (COPD GOLD spirometry grade 2 or higher) and pathology of emphysema. IPF subjects had pathology of usual interstitial pneumonia (UIP) or honeycombing. Abbreviations: COPD, chronic obstructive pulmonary disease, IPF idiopathic pulmonary fibrosis, $FEV_1$ forced expiratory volume in 1 second, BMI body mass index, LAA950 percentage of lung voxels less than a threshold of -950 Hounsfield units on chest CT scan, HP hypersensitivity pneumonitis, NSCLC non-small cell lung cancer, SCLC small cell lung cancer, UIP usual interstitial pneumonia, sSNV somatic single nucleotide variant

| Outcome: FVC (L) | All (n=1,145) | | COPD (n=335) | | IPF (n=146) | |
|---|---|---|---|---|---|---|
| | Beta (95% CI) | P value | Beta (95% CI) | P value | Beta (95% CI) | P value |
| Total sSNV* | -0.24 (-0.30, -0.19) | $<2x10^{-16}$ | -0.11 (-0.20, -0.01) | 0.03 | -0.25 (-0.37,-0.12) | 0.0001 |
| Age* | -0.09(-0.14, -0.04) | $<2x10^{-16}$ | 0.02 (-0.05, 0.1) | 0.58 | 0.09 (-0.02,0.21) | 0.12 |
| Male sex | 0.35 (0.21, 0.49) | $<8x10^{-7}$ | 0.52 (0.29, 0.76) | $<2x10^{-5}$ | 0.42 (0.06, 0.78) | 0.02 |
| Race | | | | | | |
| African American | 0.50 (0.08,0.91) | 0.02 | 0.16 (-0.52, 0.83) | 0.64 | 0.03 (-1.5, 1.5) | 0.97 |
| Hispanic | 0.04 (-0.28, 0.37) | 0.79 | -0.45 (-1.1, 0.17) | 0.15 | 0.16 (-0.47, 0.78) | 0.63 |
| Other | -0.12 (-0.65,0.42) | 0.67 | 0.30 (-0.67, 1.27) | 0.54 | -0.002 (-0.8, 0.8) | 0.99 |
| Height (cm)* | 0.41 (0.34, 0.48) | $<2x10^{-16}$ | 0.43 (0.31, 0.55) | $<2x10^{-12}$ | 0.28 (0.12, 0.44) | 0.001 |
| Smoking (PY)* | -0.02(-0.07, 0.03) | 0.35 | 0.02 (-0.05, 0.1) | 0.53 | 0.11 (0, 0.23) | 0.05 |

**Supplementary Table 3. Lung function and somatic mutational burden.**
Lung function (FVC (L)) is associated with lung somatic mutational burden. Multivariable linear regression. *Continuous variables are standardized. FVC forced vital capacity. COPD chronic obstructive pulmonary disease, IPF idiopathic pulmonary fibrosis. sSNV somatic single nucleotide variant, PY pack-years

| Cell type | Marker genes |
|---|---|
| Basal | *KRT5, KRT15, MIR305HG, KRT6A, LGALS7B* |
| Aberrant basaloid | *KRT17, TP63, SOX4, CDKN2A, ITGAV, FN1, CDH2, PRSS2* |

**Supplementary Table 4. Marker genes used for marker-based deconvolution of aberrant basaloid cells.**

| Outcome: Total sSNV | All (n=1,143) | | COPD (n=333) | | IPF (n=146) | |
|---|---|---|---|---|---|---|
| | Beta (95% CI) | P value | Beta (95% CI) | P value | Beta (95% CI) | P value |
| Airway:Alveolar* | 0.15 (0.13, 0.17) | $<2x10^{-16}$ | 0.04 (0.007, 0.07) | 0.016 | 0.20 (0.16, 0.25) | $<1 \times 10^{-15}$ |
| FVC (L)* | -0.05 (-0.07, -0.03) | $<3x10^{-7}$ | -0.05 (-0.09, -0.01) | 0.012 | -0.07 (-0.11, -0.02) | 0.005 |
| Age* | -0.006 (-0.02, 0.01) | 0.5 | -0.01 (-0.04, 0.02) | 0.37 | 0.002(-0.04, 0.04) | 0.92 |
| Male sex | -0.01 (-0.05, 0.03) | 0.71 | -0.03 (-0.11, 0.05) | 0.42 | -0.01 (-0.11, 0.09) | 0.8 |
| Race | | | | | | |
| African American | 1.06 (0.94, 1.18) | $<2x10^{-16}$ | 1.12 (0.89, 1.35) | $<2x10^{-16}$ | 0.84 (0.37, 1.3) | 0.001 |
| Hispanic | 0.18 (0.07, 0.29) | 0.001 | 0.07 (-0.18, 0.32) | 0.58 | 0.20 (-0.01, 0.41) | 0.06 |
| Other | 0.15 (-0.03, 0.33) | 0.1 | -0.28 (-0.68, 0.12) | 0.18 | 0.05 (-0.24, 0.33) | 0.75 |
| Smoking (PY)* | -0.01(-0.03, 0.005) | 0.18 | -0.01 (-0.04, 0.02) | 0.45 | 0.02 (-0.02, 0.06) | 0.34 |
| Somatic coverage* | -0.006 (-0.02, 0.01) | 0.47 | -0.006 (-0.04, 0.03) | 0.72 | -0.03 (-0.07, 0.01) | 0.13 |

**Supplementary Table 5. Predictors of lung somatic mutational burden.**
Lung somatic mutation burden is associated with lung function (FVC) and airway to alveolar epithelial ratio. The number of total sSNV was log-transformed. *Continuous variables are standardized. FVC forced vital capacity. COPD chronic obstructive pulmonary disease, IPF idiopathic pulmonary fibrosis. sSNV somatic single nucleotide variant, PY pack-year

| Top 20 FLAGS |
|---|
| *TTN, MUC16, OBSCN, AHNAK2, SYNE1, FLG, MUC5B, DNAH17, PLEC, DST, SYNE2, NEB, HSPG2, LAMA5, AHNAK, HMCN1, USH2A, DNAH11, MACF1, MUC17* |

**Supplementary Table 6. Top 20 frequently mutated genes used for sensitivity analysis**
