## Supplementary Figures for "Somatic mutations in chronic lung disease are associated with reduced lung function"

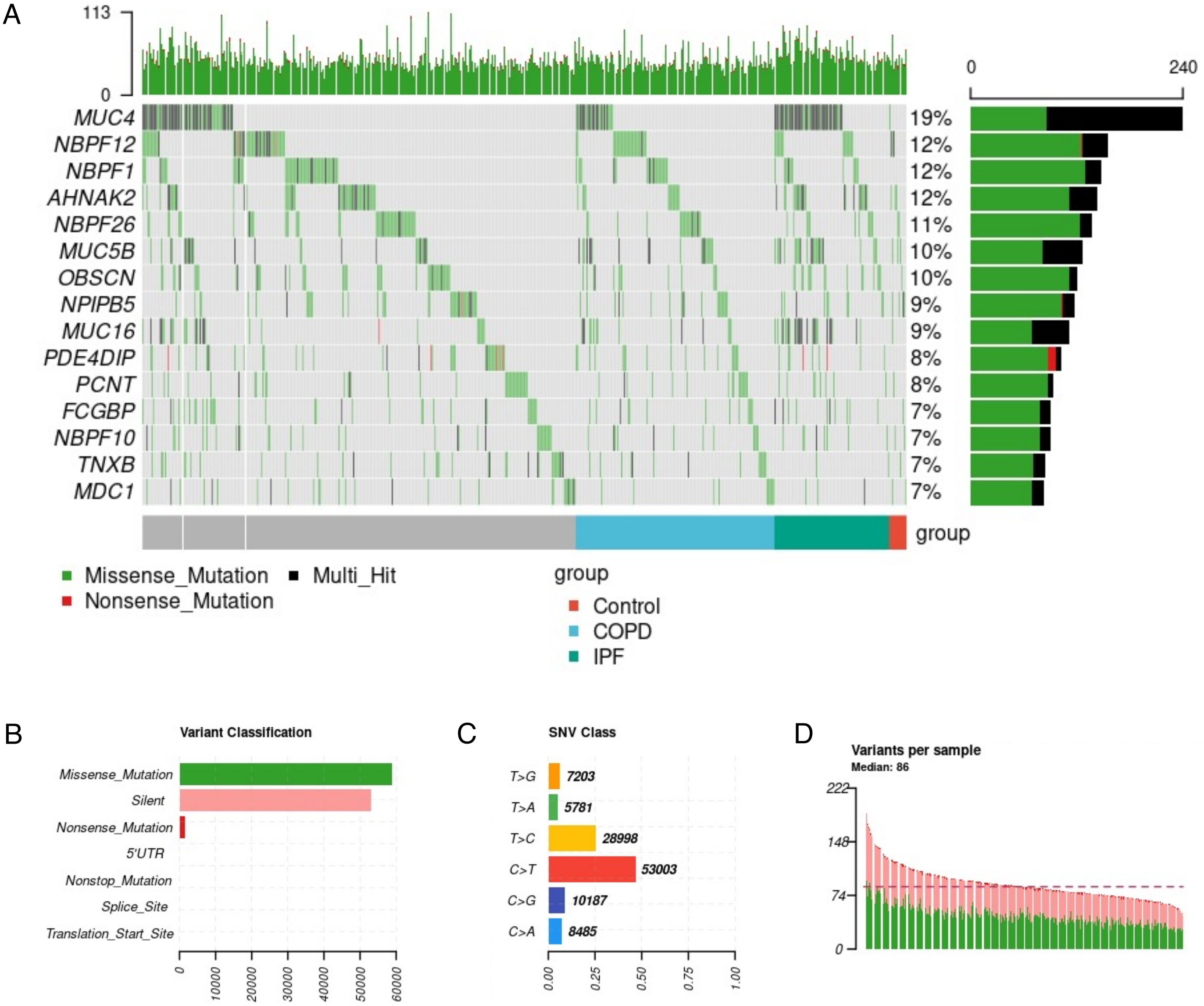


**Extended Data Figure 1. Summary of somatic mutations from lung tissue**
Samples excluding 13 cancer samples. A. Oncoplot depicting nonsynonymous mutations B. Frequency of variant types C. variant class and D. variant distribution per sample. Plot generated by maftools

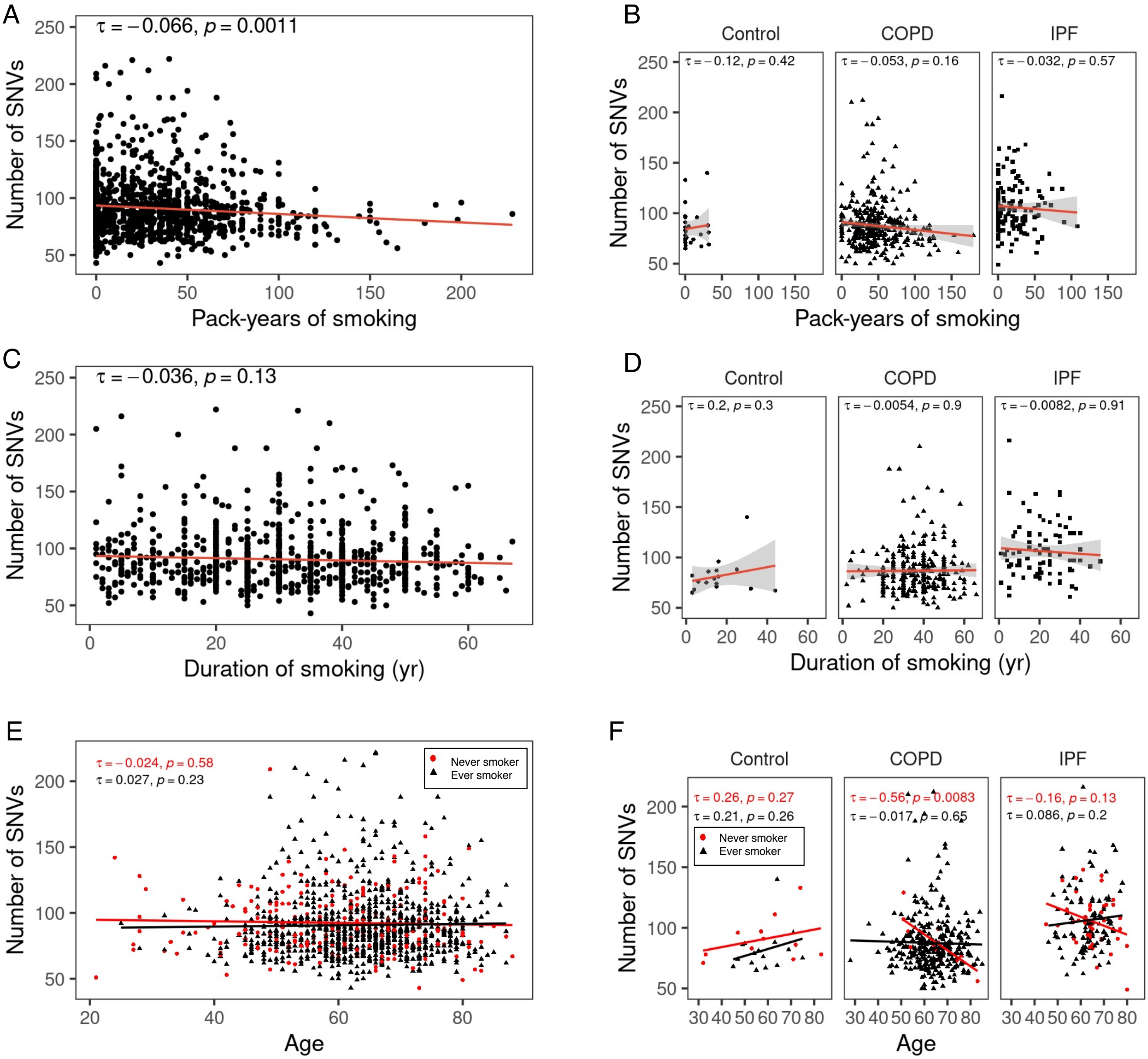


**Extended Data Figure 2. Somatic mutational burden is not significantly associated with smoking history**

A. Somatic mutational burden is weakly associated with smoking history in overall population. B. Somatic mutational burden is not associated with smoking history in disease subgroups. C Somatic mutational burden is no associated duration of smoking (years smoked) in overall subjects and D in disease subgroups. E. There is no interaction between smoking status and somatic mutational burden across age in all groups and F disease subgroups. Nonsmoker COPD subjects have lower SNVs with age. Red circle denotes never smoker, black triangle depicts ever smoker. Kendall rank correlation coefficient shown. Abbreviations: SNV single nucleotide variant, COPD chronic obstructive pulmonary disease, IPF idiopathic pulmonary fibrosis

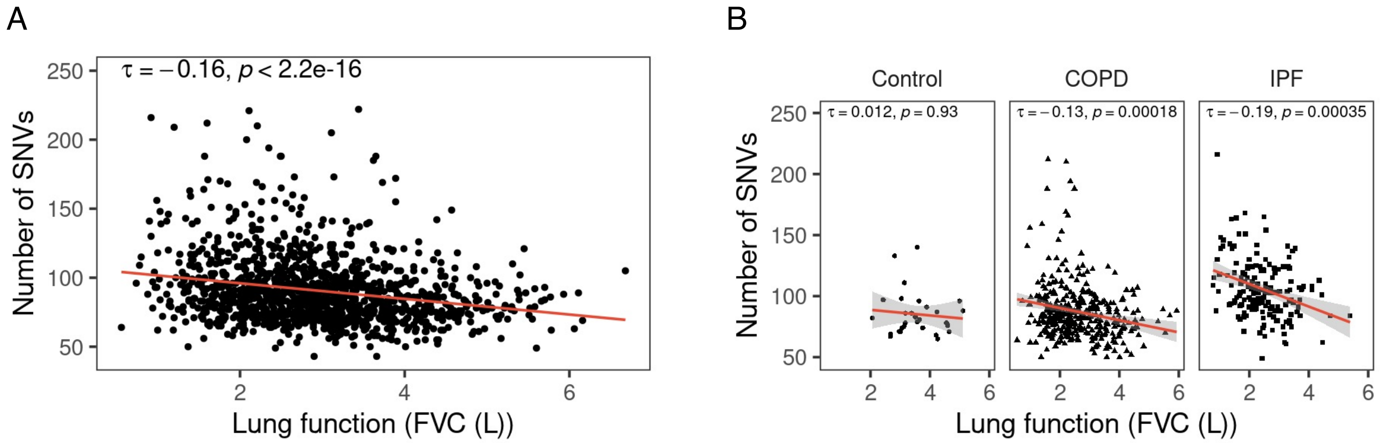


**Extended Data Fig 3. Mutational burden and lung function (FVC) relationship.** Number of somatic SNVs are inversely associated with FVC in A. all samples and B. COPD and IPF. Kendall rank correlation coefficient shown. Abbreviations: SNV single nucleotide variant, COPD chronic obstructive pulmonary disease, IPF idiopathic pulmonary fibrosis, FVC forced vital capacity

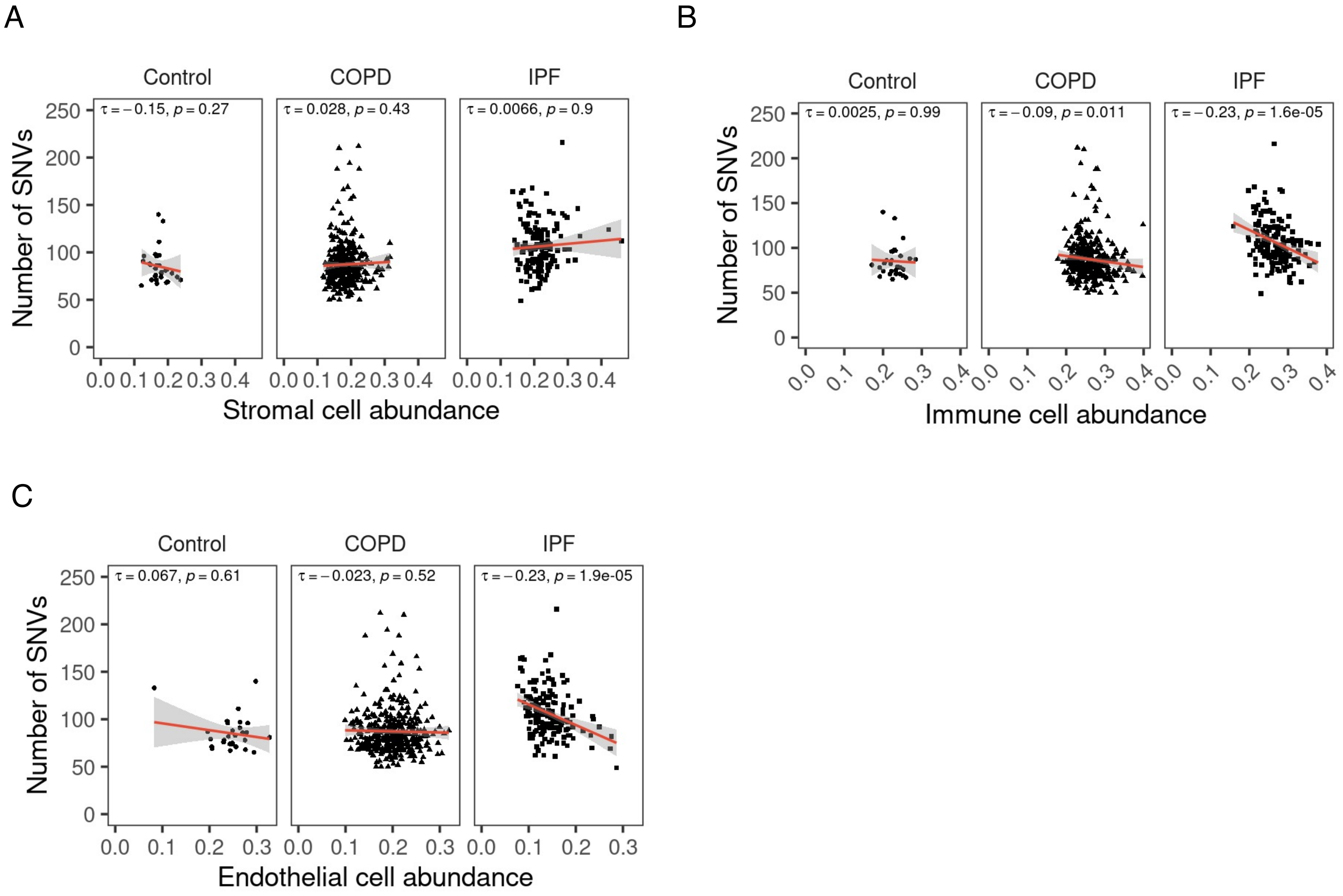


**Extended Data Fig 4. Mutational burden and cell type association is cell type specific.** Mutational burden and association with A. Stromal cells B. Immune cell and C. Endothelial cell abundance inferred from deconvolution of the RNAseq data using Bisque. Abbreviations: SNV single nucleotide variant, COPD chronic obstructive pulmonary disease, IPF idiopathic pulmonary fibrosis. Kendall rank correlation coefficient shown.

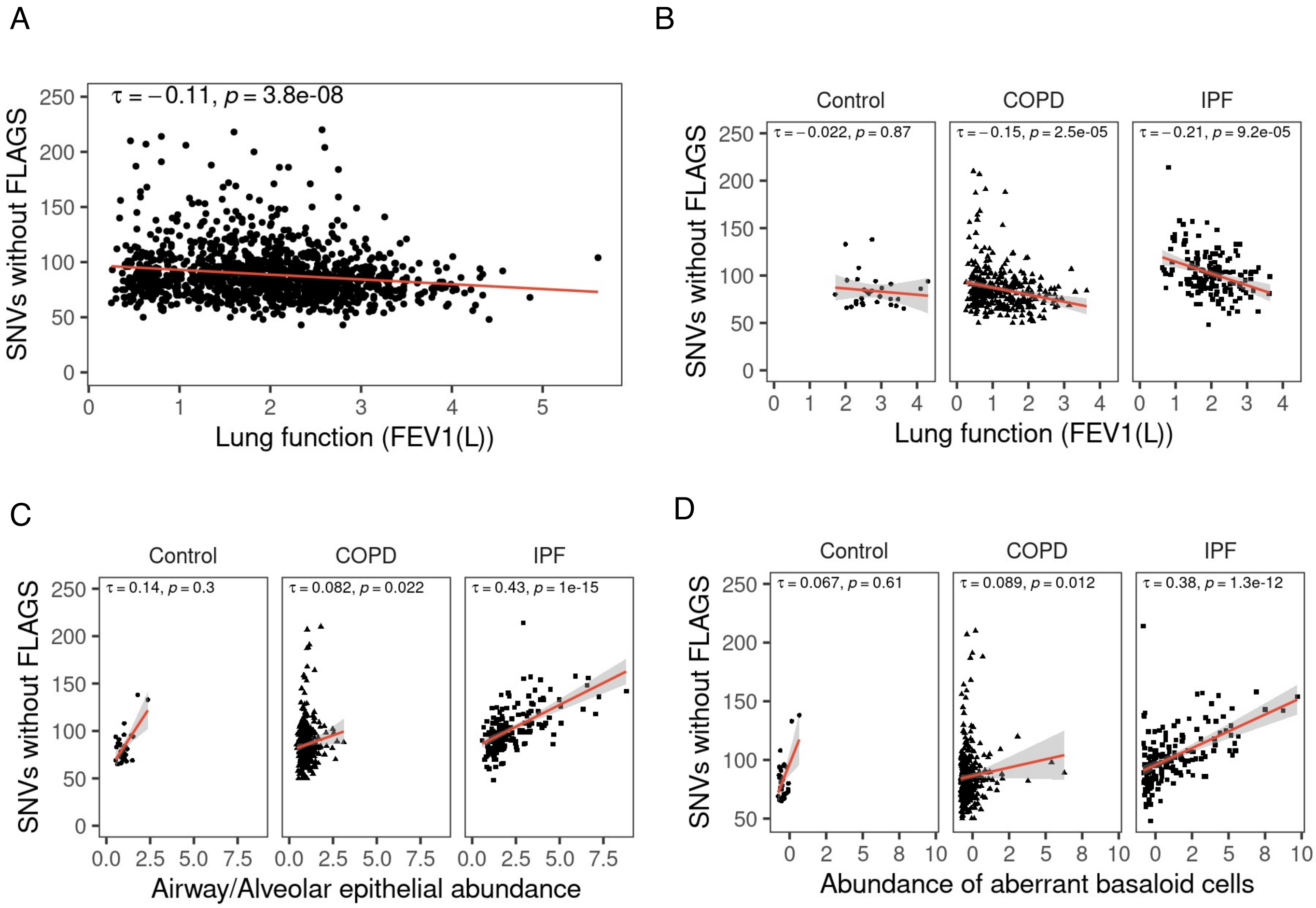


**Extended Data Fig 5. Mutational burden after excluding top 20 FLAGS genes.** Mutational burden after excluding top 20 FLAGS genes (*TTN, MUC16, OBSCN, AHNAK2, SYNE1, FLG, MUC5B, DNAH17, PLEC, DST, SYNE2, NEB, HSPG2, LAMA5, AHNAK, HMCN1, USH2A, DNAH11, MACF1, MUC17*). Abbreviations: SNV single nucleotide variant, COPD chronic obstructive pulmonary disease, IPF idiopathic pulmonary fibrosis. Kendall rank correlation coefficient shown.

A

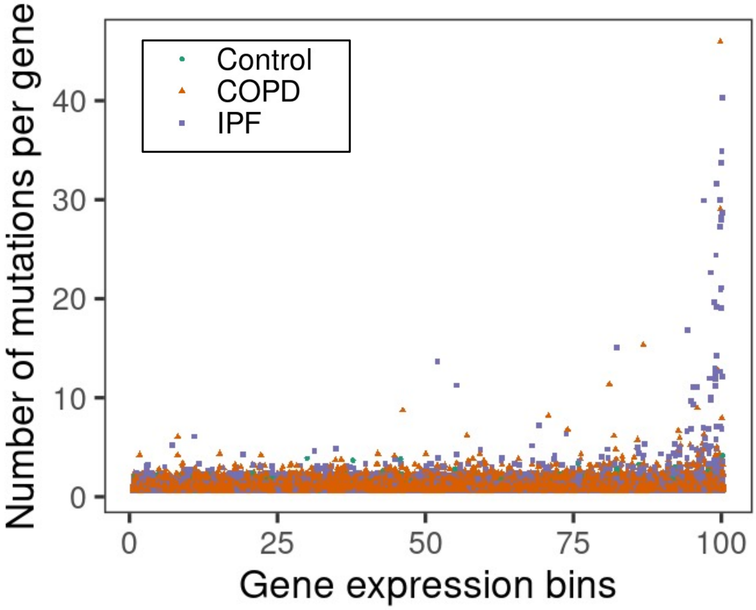


B

| Bin | Comparison | Difference (95% CI) | adj. P |
|---|---|---|---|
| 99 | IPF- normal | 1.24 (-0.01, 2.48) | 0.05 |
| 99 | IPF-COPD | 1.18 (0.59, 1.77) | 0.00001 |

**Extended Data Fig 6. Number of mutations per gene by length normalized gene expression categories.**

A. Genes are divided into 100 bins based on length scaled TPM. B. Within each gene expression bin, differences in number of mutations per gene by disease group was tested using ANOVA with Bonferroni adjustment. In genes with high expression (bin 99) IPF samples had higher number of mutation per gene compared to normal and COPD.

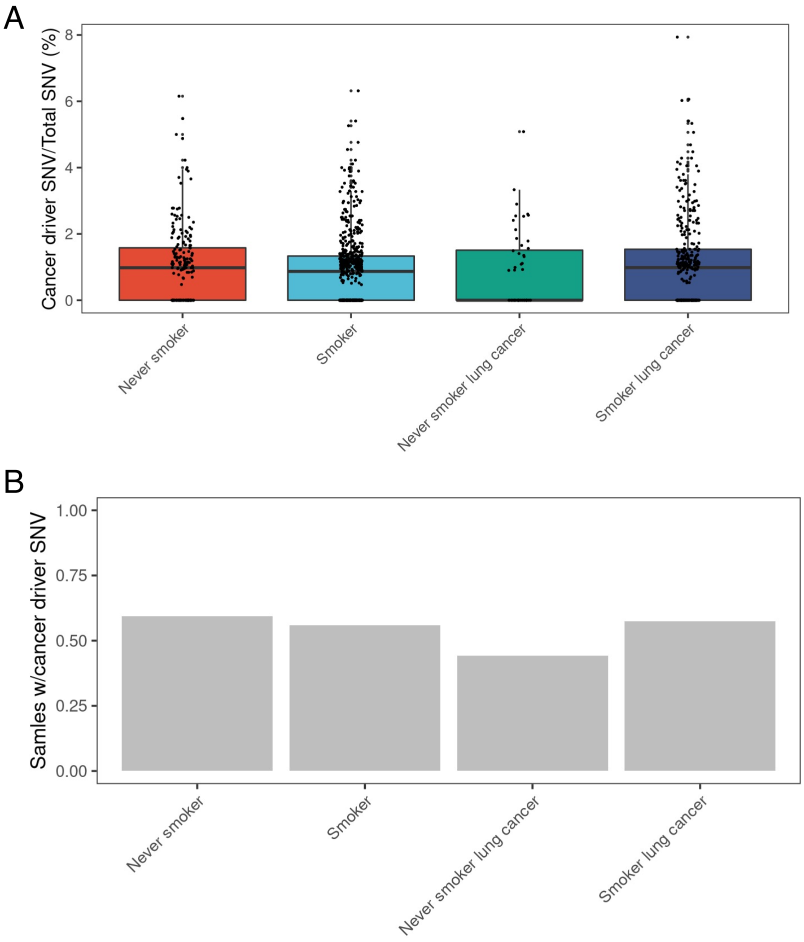


**Extended Data Fig 7. Cancer driver gene SNVs at sample and population level.**
A. Samples stratified by smoking and lung cancer history. Proportion of cancer driver gene SNV (number of caner driver gene SNV divided by total SNV of sample) at individual sample level. B at population level. subjects with any cancer driver gene SNV divided by total number of subjects.

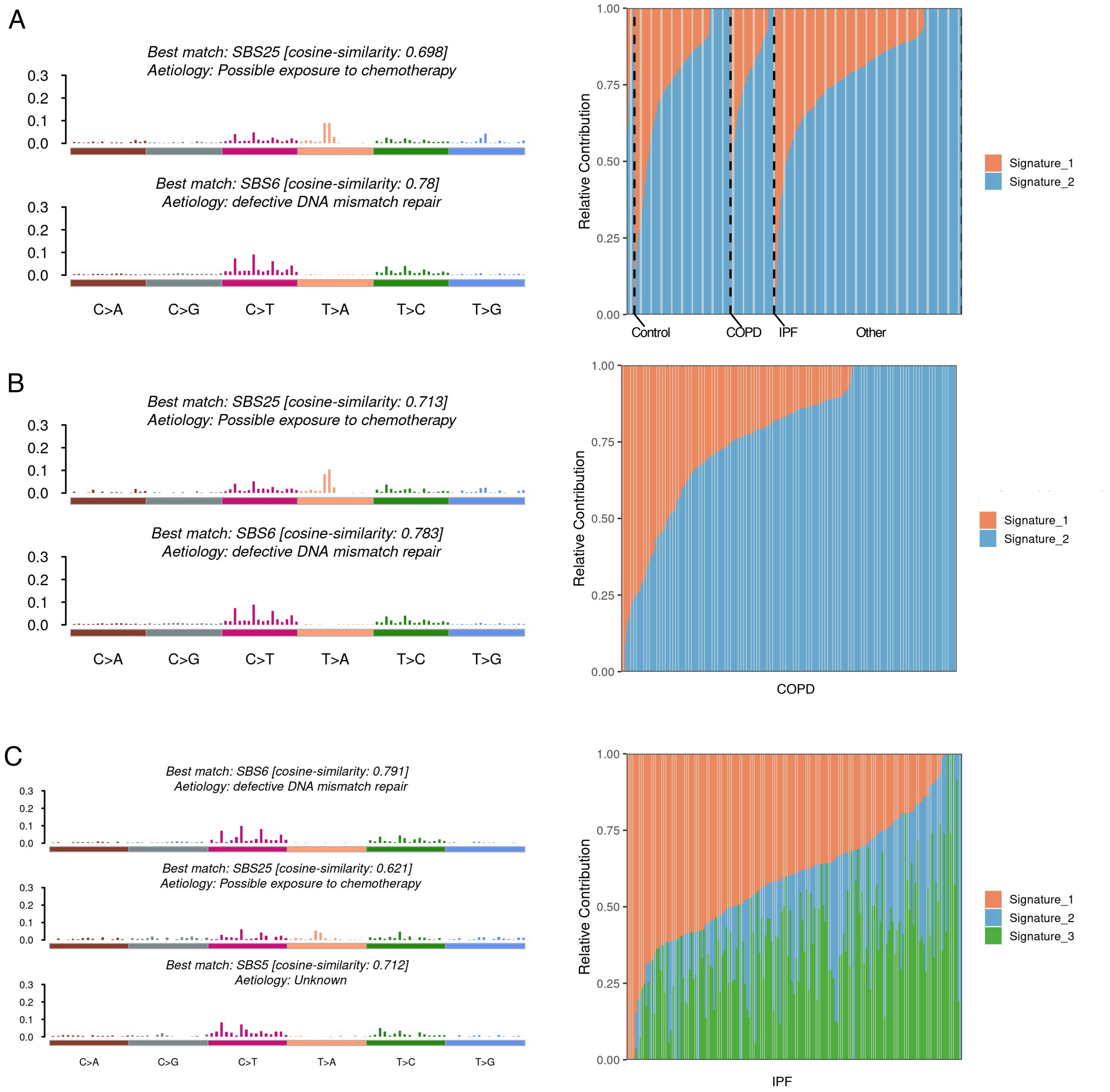


**Extended Data Fig 8. Somatic mutational signatures in chronic lung disease**
Single base substitution (SBS) signatures found in A. all subjects, B. Chronic obstructive pulmonary disease (COPD) subjects, C. Idiopathic pulmonary fibrosis (IPF) subjects. Left panel showing chronic lung disease mutations decomposed into COSMIC signatures (SBS 25, 6, and 5). Right panel showing relative contribution of mutational signatures in individual samples.
